# One test is enough: Japan’s two-test rule for HIV treatment subsidies delays antiretroviral therapy without changing eligibility

**DOI:** 10.64898/2026.09.28.26364228

**Authors:** Toshibumi Taniguchi, Takashi Muramatsu, Rumi Minami, Hideta Nakamura, Eisuke Adachi, Ryoko Sekiya, Kazuaki Fukushima, Junji Imamura, Yusuke Yoshino, Michinori Shirano, Yuji Hirai, Mayumi Imahashi, the J-HIV RWD Collaborative Database Team

## Abstract

**Introduction:** Japan achieves high retention and viral suppression among people on antiretroviral therapy, yet treatment starts late. Most people obtain subsidised treatment through a physical disability certificate for HIV-related immune dysfunction, which requires two blood tests at least four weeks apart. We quantified the diagnosis-to-treatment cascade, identified who waits and tested whether the second test changes certification.

**Methods:** Retrospective cohort of people newly diagnosed with HIV at 11 designated AIDS core hospitals in 2015 to 2024. We measured intervals from screening to hospital visit and to antiretroviral therapy, and emulated certification from the first test alone and from the two-test rule under every possible number of non-laboratory criteria (k = 0 to 8).

**Results:** Of 1302 people (96% men; 68% with CD4 <350 cells/µL or AIDS), 1036 (80%) followed the two-test pathway and 261 started treatment after one test, mostly because of AIDS or acute infection. People reached the hospital a median of 11 days after screening. Treatment then began a median of 70 days after the first visit in the two-test pathway and 11 days in the single-test pathway; 0.2% and 60.9% started within 14 days. Delay rose with CD4 count: 46 days when the mean count was ≤200 cells/µL, 117 days when it was >500 with a laboratory criterion (16% of the two-test pathway) and 334 days when it was >500 without one (3%; adjusted hazard ratio 0.14, 95% confidence interval 0.09 to 0.23). Medians ranged from 35 to 96 days across facilities. The second test changed little: the first CD4 count agreed with the mean of two (intraclass correlation coefficient 0.959), HIV-RNA ≥5000 copies/mL persisted in 97.6%, and certification status agreed in 99.0% for k ≥1 (92.5% for k = 0), with discordance concentrated near 500 cells/µL; the first test gave a milder grade in at most 2.9%.

**Conclusions:** The statutory second test adds at least four weeks before antiretroviral therapy in Japan without changing who is certified, and delays most the people with the highest CD4 counts. Certification on a single test, with treatment separated from the subsidy procedure, would remove a delay that has no clinical or administrative justification.

## Introduction

Antiretroviral therapy (ART) is recommended for everyone with HIV, and guidelines advise starting it promptly, within seven days of diagnosis in World Health Organization guidance [1–4]. Trials [5,6], a citywide programme [7] and systematic reviews [8–10] show that rapid ART increases treatment uptake and retention with similar or better viral suppression, and a suppressed viral load prevents transmission [11,12].

Japan has a concentrated epidemic among men who have sex with men, and its cascade performs well once people are in care: retention exceeds 95% and viral suppression among those on ART exceeds 99% [13,14]. The beginning of the cascade is weaker. About one third of new HIV/AIDS reports are already at the stage of AIDS [15], and ART starts late: only 8% of people with acute or recent infection started within two weeks in a national survey [16], and a single-centre study found a median of 42 days from first visit to ART [17]. The main reason is statutory. ART is covered by public health insurance with a 30% co-payment, and most people reduce this through a physical disability certificate for “immune dysfunction due to HIV”, which gives access to the medical care subsidy for persons with disabilities [18–21]. The laboratory requirements of the criteria, unchanged since the category was created in 1998, require the CD4 count to be the mean of two tests at least four weeks apart and each laboratory criterion (white blood cell count, haemoglobin, platelets, HIV-RNA) to be met on both tests, unless the person has an AIDS-defining illness (Supplementary Text S1). Because the subsidy is not retroactive, clinicians commonly wait for the second test and certification before starting ART.

Whether the second test serves any purpose has not been examined. Within-person variation of CD4 counts is well described [22–25], and single measurements misclassify people near a threshold of 500 [26] or 200 cells/µL [27], but no study has measured how much delay the rule adds, who bears it, or how often the two tests differ. We therefore analysed a multicentre cohort of people newly diagnosed with HIV in Japan to quantify the diagnosis-to-treatment cascade, to identify who waits, and to test whether certification from the first test alone would differ from the two-test rule.

## Methods

### Study design and setting

We conducted a retrospective multicentre cohort study at 11 designated AIDS core hospitals in seven prefectures from Sendai to Okinawa that participate in the J-HIV RWD Collaborative Database Team. Each hospital entered consecutive people newly diagnosed with HIV into a web-based registration system (UMIN Internet Data and Information Center for Medical Research). We followed the STROBE statement and the Guidelines for Reporting Reliability and Agreement Studies [28,29].

### Participants

Eligible participants were people aged 13 years or older (the criteria for children differ) who were newly diagnosed with HIV, were not on ART at referral, and had their first blood test at the participating hospital between 1 January 2015 and 31 December 2024. Records were entered between October 2025 and April 2026 and included follow-up to the last recorded visit.

### Data collection

From medical records, each hospital recorded demographics, transmission category, dates of HIV screening and confirmatory tests, the dates and results of the first blood test at the hospital and of the second test used for certification (CD4 count, HIV-RNA, white blood cell count, haemoglobin, platelets), AIDS-defining illnesses, the date and regimen of ART, the reason for starting ART after one test, dates of HIV-RNA suppression, CD4 counts at 12 to 60 months after ART, and the end of follow-up. Data were checked for implausible values and date errors.

### Definitions

The date of the first blood test at the hospital was taken as the date of the first visit. Pathway was classified from the recorded reason for ART initiation: the two-test pathway comprised people recorded as starting ART after two tests who had a valid pair of CD4 counts (ART on the day of the second draw was allowed), plus 10 people who had not started ART but had a valid pair at least 28 days apart; the single-test pathway comprised people recorded as starting ART after one test (four also had a second test drawn before or on the day of ART). Late presentation was a first CD4 count <350 cells/µL or an AIDS-defining illness. The interval between tests was not restricted in the main analysis (32 pairs were <28 days apart); a sensitivity analysis used pairs 28 to 90 days apart.

We emulated the certification criteria [18] in two ways. The first-test judgement used the first CD4 count and the laboratory criteria met at the first test; the two-test rule used the mean of the two counts and the criteria met on both tests. The eight non-laboratory criteria (items e to l, symptoms and daily-life restrictions; Text S1) were not available in medical records. To isolate the contribution of repeat laboratory testing, we held the assumed number of non-laboratory criteria, k, constant between the two judgements and repeated the comparison for every k from 0 to 8. The primary comparison assumed k ≥1, because the certification guidance counts strict medication management as a daily-life restriction [19], which applies to essentially everyone about to start lifelong ART; k = 0 is a formal sensitivity scenario. Under the grading rules (Text S1), certification status therefore differs between judgements only through the CD4 count and the laboratory criteria.

To describe who waits, we divided the two-test pathway into three groups under the two-test rule: mean CD4 count ≤500 cells/µL (certifiable when k ≥1); >500 cells/µL with at least one persistent laboratory criterion; and >500 cells/µL with none (not certifiable at any k).

### Outcomes

The primary outcomes were days from the screening test to the first visit, from the first visit to ART and from the second test to ART; rapid ART, defined as ART within 14 days of the first visit; and the proportions who had started ART by 90, 180 and 365 days. The agreement outcomes were the intraclass correlation coefficient (ICC), bias and 95% limits of agreement between the first CD4 count and the mean of two counts and between the first and second counts; agreement in CD4 categories (≤200, 201 to 500, >500 cells/µL) with Cohen’s kappa; persistence of each laboratory criterion at the second test; and agreement in certification status and grade between the two judgements, with the one-sided 95% upper confidence bound (Wilson) for the proportion in which the first test gave a milder grade compared against a 5% reference threshold. Secondary outcomes were ART regimens, time to HIV-RNA <200 copies/mL and CD4 counts 12 and 24 months after ART.

### Statistical analysis

Time to ART was analysed with Kaplan-Meier estimates, censoring people who had not started ART at the last follow-up date, and compared with Mann-Whitney and Kruskal-Wallis tests. Cox models stratified by facility (facilities of fewer than 20 people pooled) estimated hazard ratios for ART initiation adjusted for age, sex, transmission category and year of first visit. We calculated ICC(2,1) for absolute agreement with bootstrap 95% confidence intervals (1000 resamples) and Bland-Altman limits of agreement [30,31] and a within-person coefficient of variation (Table 4). Persistence was summarised with Wilson 95% confidence intervals. Missing laboratory values at the first test (four values in two people) were treated as not meeting the criterion. Logistic regression estimated the odds of discordance between judgements per 50 cells/µL distance of the first CD4 count from 500 cells/µL. Time to viral suppression was analysed with Kaplan-Meier estimates and Cox models stratified by facility and adjusted for pre-treatment HIV-RNA and CD4 count, age, sex, AIDS-defining illness, pathway, year and anchor drug, with a model stratified by HIV-RNA category because Schoenfeld residuals showed non-proportional hazards for HIV-RNA; linear regression of CD4 counts at 12 and 24 months included the log-transformed time from first visit to ART (Tables S3A to S3D). Post-ART outcomes excluded one hospital that did not record them. Analyses used Python 3.10.

### Ethics

The study was approved by the Ethics Review Committee for Observational Research of Chiba University Hospital (reference HK202508-12) as the central committee, with the consent requirement waived under an opt-out procedure, and was authorised by each participating hospital.

## Results

### Participants and presentation

Overall, 1302 people were included (Figure 1); the median age was 36 years, 96.2% were men and 85.6% acquired HIV through sex between men (Table 1). Presentation was late: the median first CD4 count was 248 cells/µL (IQR 73 to 400), 42.2% had a count ≤200 cells/µL, 26.2% had an AIDS-defining illness and 68.3% met the definition of late presentation. In total, 1036 people (79.6%) followed the two-test pathway and 261 (20.0%) started ART after a single test, most often for AIDS (n = 212); five had neither a second test nor ART. Overall, 1287 people (98.8%) started ART, 98% with an integrase inhibitor-based regimen (Table S2); ART was not recorded in 15 people, eight lost to follow-up and seven transferred to another hospital.

**Table 1.**
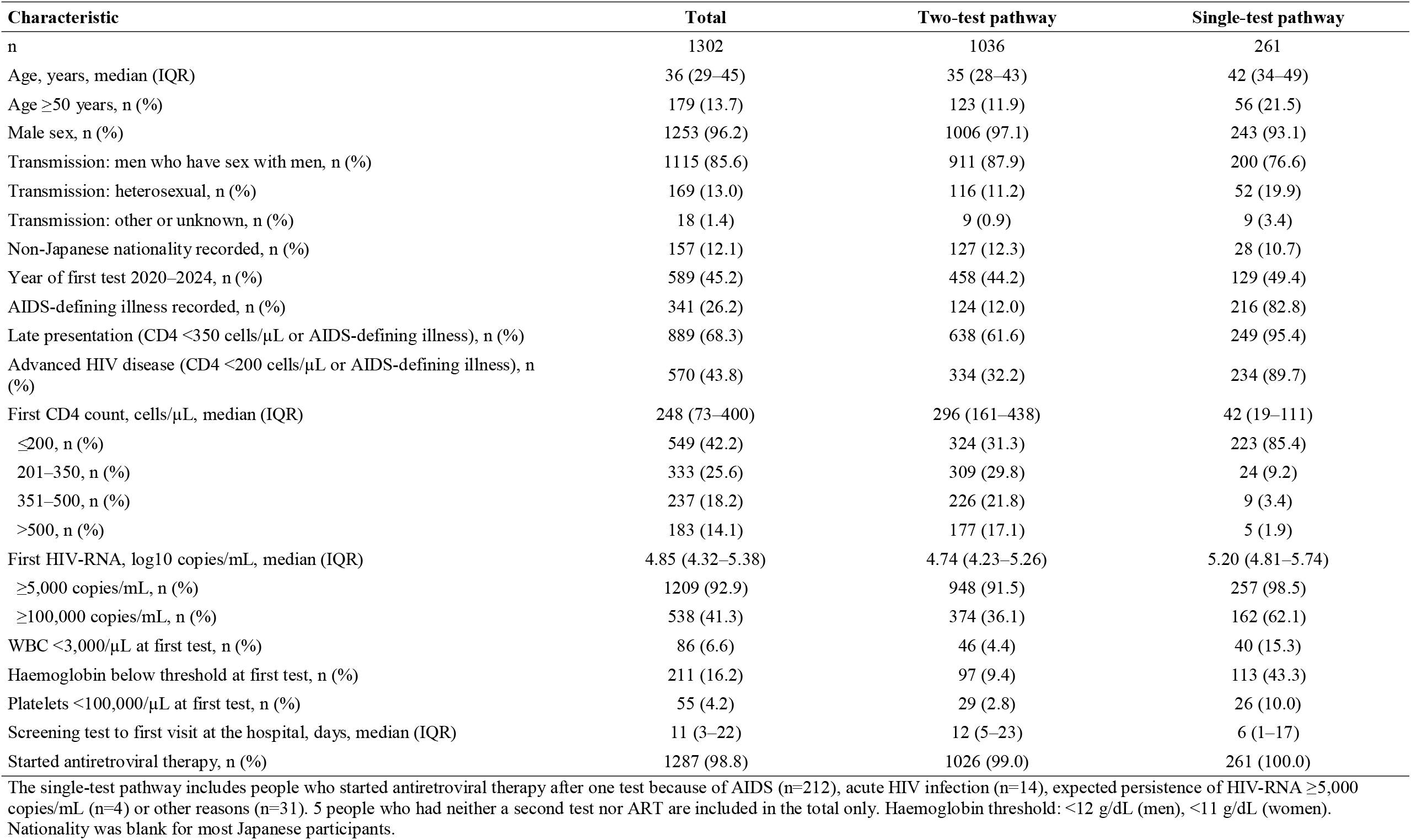
Characteristics of people newly diagnosed with HIV at 11 designated AIDS core hospitals in Japan, 2015 to 2024.

| Characteristic | Total | Two-test pathway | Single-test pathway |
| --- | --- | --- | --- |
| n | 1302 | 1036 | 261 |
| Age, years, median (IQR) | 36 (29–45) | 35 (28–43) | 42 (34–49) |
| Age ≥50 years, n (%) | 179 (13.7) | 123 (11.9) | 56 (21.5) |
| Male sex, n (%) | 1253 (96.2) | 1006 (97.1) | 243 (93.1) |
| Transmission: men who have sex with men, n (%) | 1115 (85.6) | 911 (87.9) | 200 (76.6) |
| Transmission: heterosexual, n (%) | 169 (13.0) | 116 (11.2) | 52 (19.9) |
| Transmission: other or unknown, n (%) | 18 (1.4) | 9 (0.9) | 9 (3.4) |
| Non-Japanese nationality recorded, n (%) | 157 (12.1) | 127 (12.3) | 28 (10.7) |
| Year of first test 2020–2024, n (%) | 589 (45.2) | 458 (44.2) | 129 (49.4) |
| AIDS-defining illness recorded, n (%) | 341 (26.2) | 124 (12.0) | 216 (82.8) |
| Late presentation (CD4 <350 cells/μL or AIDS-defining illness), n (%) | 889 (68.3) | 638 (61.6) | 249 (95.4) |
| Advanced HIV disease (CD4 <200 cells/μL or AIDS-defining illness), n (%) | 570 (43.8) | 334 (32.2) | 234 (89.7) |
| First CD4 count, cells/μL, median (IQR) | 248 (73–400) | 296 (161–438) | 42 (19–111) |
| ≤200, n (%) | 549 (42.2) | 324 (31.3) | 223 (85.4) |
| 201–350, n (%) | 333 (25.6) | 309 (29.8) | 24 (9.2) |
| 351–500, n (%) | 237 (18.2) | 226 (21.8) | 9 (3.4) |
| >500, n (%) | 183 (14.1) | 177 (17.1) | 5 (1.9) |
| First HIV-RNA, log10 copies/mL, median (IQR) | 4.85 (4.32–5.38) | 4.74 (4.23–5.26) | 5.20 (4.81–5.74) |
| ≥5,000 copies/mL, n (%) | 1209 (92.9) | 948 (91.5) | 257 (98.5) |
| ≥100,000 copies/mL, n (%) | 538 (41.3) | 374 (36.1) | 162 (62.1) |
| WBC <3,000/μL at first test, n (%) | 86 (6.6) | 46 (4.4) | 40 (15.3) |
| Haemoglobin below threshold at first test, n (%) | 211 (16.2) | 97 (9.4) | 113 (43.3) |
| Platelets <100,000/μL at first test, n (%) | 55 (4.2) | 29 (2.8) | 26 (10.0) |
| Screening test to first visit at the hospital, days, median (IQR) | 11 (3–22) | 12 (5–23) | 6 (1–17) |
| Started antiretroviral therapy, n (%) | 1287 (98.8) | 1026 (99.0) | 261 (100.0) |

The single-test pathway includes people who started antiretroviral therapy after one test because of AIDS (n=212), acute HIV infection (n=14), expected persistence of HIV-RNA ≥5,000 copies/mL (n=4) or other reasons (n=31). 5 people who had neither a second test nor ART are included in the total only. Haemoglobin threshold: <12 g/dL (men), <11 g/dL (women). Nationality was blank for most Japanese participants.

**Figure 1.**
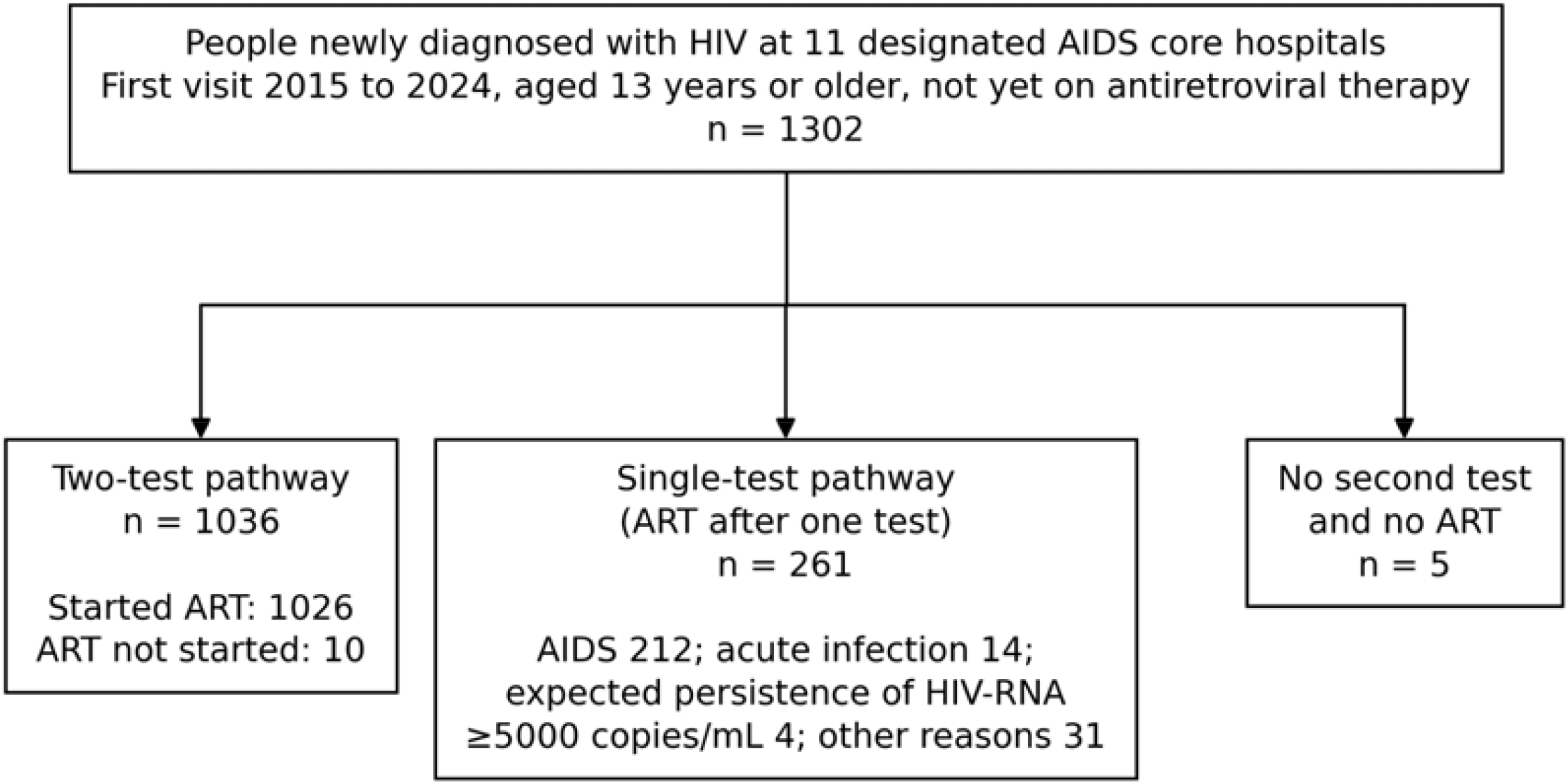
Flow of participants. People newly diagnosed with HIV at the 11 designated AIDS core hospitals who met the inclusion criteria, and their assignment to the two-test and single-test pathways. ART, antiretroviral therapy.

### The diagnosis-to-treatment cascade

People reached the hospital a median of 11 days (IQR 3 to 22) after screening (n = 1295 with both dates), and 16.5% took more than 30 days. From the first visit, ART began after a median of 56 days (IQR 29 to 92): 70 days (IQR 42 to 106) in the two-test pathway and 11 days (IQR 4 to 21) in the single-test pathway (p < 0.001; Table 2, Figure 2A). In the two-test pathway, a median of 29 days (IQR 28 to 35) elapsed between the tests and a further 35 days (IQR 9 to 68) between the second test and ART. Rapid ART was achieved by 0.2% of the two-test pathway and 60.9% of the single-test pathway; 65.9% and 98.5% had started by 90 days. Intervals were similar in men and women and shortened only modestly over time (Table S5).

**Table 2.** Time to antiretroviral therapy by testing pathway and by certifiability under the two-test rule.

| Group | n | Started ART, n | Screening to ART, days, median (IQR) | First test to ART, days, median (IQR) | Second test to ART, days, median (IQR) | ART within 14 days, % | ART within 90 days, % | ART within 180 days, % | ART within 365 days, % | Adjusted HR for ART initiation (95% CI) |
| --- | --- | --- | --- | --- | --- | --- | --- | --- | --- | --- |
| Two-test pathway | 1036 | 1026 | 87 (56–132) | 70 (42–106) | 35 (9–68) | 0.2 | 65.9 | 89.2 | 94.0 | 0.12 (0.09–0.15) |
| Single-test pathway | 261 | 261 | 22 (12–33) | 11 (4–21) | NA | 60.9 | 98.5 | 98.5 | 98.5 | 1 (reference) |
| AIDS at diagnosis | 212 | 212 | 21 (13–30) | 12 (5–21) | NA | 59.9 | 98.1 | 98.1 | 98.1 |  |
| Acute HIV infection | 14 | 14 | 23 (8–33) | 9 (2–20) | NA | 64.3 | 100.0 | 100.0 | 100.0 |  |
| Other reasons | 35 | 35 | 30 (14–44) | 6 (0–18) | NA | 65.7 | 100.0 | 100.0 | 100.0 |  |
| Two-test pathway: mean CD4 ≤500 cells/μL (certifiable) | 839 | 836 | 78 (52–114) | 63 (40–91) | 30 (7–56) | 0.2 | 74.8 | 93.9 | 97.4 | 1 (reference) |
| Two-test pathway: mean CD4 >500 with ≥1 persistent laboratory criterion | 166 | 162 | 138 (92–312) | 117 (70–182) | 72 (42–126) | 0.0 | 30.9 | 74.3 | 84.9 | 0.46 (0.39–0.56) |
| Two-test pathway: mean CD4 >500 without laboratory criteria (not certifiable) | 31 | 28 | 394 (131–807) | 334 (115–708) | 270 (74–639) | 0.0 | 12.9 | 39.7 | 50.3 | 0.14 (0.09–0.23) |
| Mean CD4 ≤200 | 297 | 296 | 57 (39–87) | 46 (31–69) | 11 (0–35) | 0.3 | 88.1 | 97.6 | 99.3 |  |
| Mean CD4 201–350 | 280 | 279 | 82 (60–115) | 67 (44–90) | 35 (14–55) | 0.0 | 74.9 | 95.3 | 97.1 |  |
| Mean CD4 351–500 | 262 | 261 | 97 (73–148) | 78 (56–114) | 47 (21–77) | 0.4 | 59.7 | 88.1 | 95.4 |  |

HR, hazard ratio for ART initiation from Cox models stratified by facility and adjusted for age, sex, MSM transmission and year of first test (pathway model additionally adjusted for AIDS-defining illness; facilities with <20 participants pooled). HR <1 indicates slower initiation. Medians (IQR) are calculated among people who started ART; percentages are Kaplan–Meier estimates that include people who had not started ART, censored at the last follow-up date. Laboratory criteria: WBC <3,000/μL, haemoglobin <12 (men)/<11 (women) g/dL, platelets <100,000/μL or HIV-RNA ≥5,000 copies/mL, each on both tests (persistent). Of the 166 people with a mean CD4 >500 cells/μL and a persistent laboratory criterion, 163 met HIV-RNA ≥5,000 copies/mL alone and could be certified only with a non-laboratory criterion; three met two laboratory criteria.

**Figure 2.**
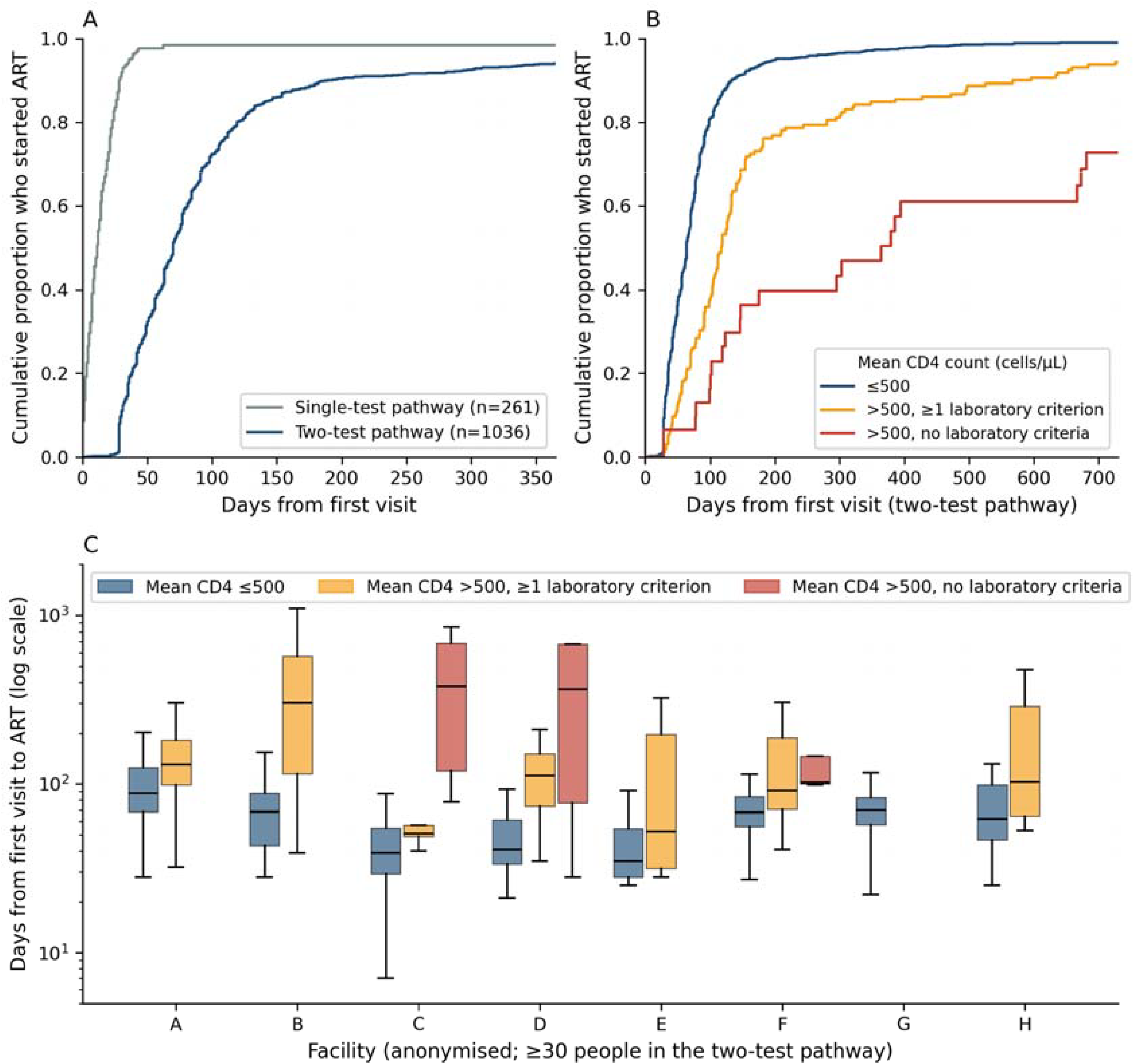
Time from the first hospital visit to antiretroviral therapy. (A) Kaplan-Meier estimates of the cumulative proportion who started antiretroviral therapy (ART) in the two-test and single-test pathways. (B) The two-test pathway by certifiability under the two-test rule: mean CD4 count ≤500 cells/µL (n = 839, certifiable), mean count >500 cells/µL with at least one laboratory criterion met on both tests (n = 166) and mean count >500 cells/µL without a persistent laboratory criterion (n = 31, not certifiable). (C) Days from the first visit to ART by facility (anonymised, facilities with at least 30 people in the two-test pathway) for the same three groups; boxes show the median and interquartile range (groups with fewer than five people are omitted).

### Who waits

Within the two-test pathway, 839 people (81.0%) had a mean CD4 count ≤500 cells/µL; 166 (16.0%, 95% CI 13.9 to 18.4) had a mean count >500 cells/µL with a persistent laboratory criterion, HIV-RNA ≥5000 copies/mL alone in 163 (certifiable only with a non-laboratory criterion) and two criteria in three; and 31 (3.0%, 95% CI 2.1 to 4.2) had a mean count >500 cells/µL with no persistent laboratory criterion and could not be certified. Time to ART rose across these groups: 63 days (IQR 40 to 91), 117 days (IQR 70 to 182) and 334 days (IQR 115 to 708; p < 0.001), and within the certifiable group from 46 days when the mean CD4 count was ≤200 cells/µL to 78 days when it was 351 to 500 cells/µL (Table 2, Figure 2B). By one year, 97.4%, 84.9% and 50.3% had started ART; three of the 31 people who could not be certified had not started at the end of follow-up. Adjusted hazard ratios were 0.46 (95% CI 0.39 to 0.56) and 0.14 (95% CI 0.09 to 0.23) for the two groups with a mean count >500 cells/µL, relative to the group with a mean count ≤500 cells/µL. Conversely, 42 people (4.1%), 33 at one facility, had an AIDS-defining illness recorded between the two tests (median first CD4 count 26 cells/µL) yet followed the two-test procedure although the AIDS exemption applied.

Facilities differed markedly (p < 0.001; Table 3, Figure 2C). Among the eight facilities with at least 30 people in the two-test pathway, median time from the first visit to ART ranged from 35 to 96 days, and from the second test to ART from 0 days, at a facility that started ART on the day of the second draw in 57% of people, to 62 days. For people with a mean CD4 count >500 cells/µL and HIV-RNA ≥5000 copies/mL, the median time to ART was 51 and 52 days at two facilities but 131 and 301 days at two others.

**Table 3.** Testing pathway and time to antiretroviral therapy by facility (anonymised)

| Facility | n | Years of first test | Two-test pathway, n (%) | First to second test, days, median (IQR) | Second test to ART, days, median (IQR) | ART on the day of the second test, % | First test to ART, days, median (IQR), two-test pathway | First test to ART, days, median (IQR), mean CD4 >500 with laboratory criteria | First test to ART, days, median (IQR), all |
| --- | --- | --- | --- | --- | --- | --- | --- | --- | --- |
| A | 372 | 2015–2024 | 331 (89.0) | 31 (28–35) | 62 (40–95) | 3.0 | 96 (72–138) | 131 (98–182) | 91 (63–133) |
| B | 217 | 2018–2020 | 153 (70.5) | 29 (28–35) | 39 (10–57) | 17.0 | 70 (44–97) | 301 (115–571) | 50 (17–84) |
| C | 168 | 2019–2024 | 152 (90.5) | 28 (28–32) | 14 (1–27) | 23.0 | 42 (31–56) | 51 (49–57) | 40 (29–56) |
| D | 145 | 2015–2024 | 65 (44.8) | 28 (28–35) | 17 (1–70) | 22.6 | 50 (35–112) | 112 (74–150) | 28 (7–42) |
| E | 136 | 2015–2024 | 125 (91.9) | 30 (28–35) | 0 (0–28) | 56.9 | 35 (28–62) | 52 (32–198) | 35 (28–56) |
| F | 122 | 2015–2024 | 99 (81.1) | 30 (28–35) | 43 (28–63) | 5.1 | 75 (58–98) | 91 (71–188) | 63 (39–88) |
| G | 71 | 2020–2024 | 52 (73.2) | 29 (28–35) | 38 (28–50) | 5.8 | 71 (62–88) | (n=4) | 63 (24–81) |
| H | 51 | 2015–2024 | 41 (80.4) | 30 (28–33) | 35 (21–87) | 2.4 | 72 (50–115) | 103 (64–288) | 60 (40–110) |
| I | 11 | 2015–2017 | 11 (100.0) | 31 (28–34) | 28 (17–60) | 0.0 | 63 (45–94) | (n=4) | 63 (45–94) |
| J | 6 | 2015–2015 | 6 (100.0) | 30 (29–33) | 14 (4–27) | 33.3 | 44 (36–57) | (n=0) | 44 (36–57) |
| K | 3 | 2018–2022 | 1 (33.3) | 31 (31–31) | 0 (0–0) | 100.0 | 31 (31–31) | (n=0) | 15 (14–23) |

### Does the second test change certification?

The first CD4 count agreed closely with the mean of the two counts (r = 0.962; ICC 0.959, 95% CI 0.952 to 0.965; bias +15.9 cells/µL, limits of agreement −101 to +133; Table 4, Figure 3), as expected because the mean shares the first count. Agreement between the first and second counts was lower (ICC 0.850, 95% CI 0.825 to 0.871; limits of agreement −202 to +265 cells/µL), with a within-person coefficient of variation of about 26%. The second count was higher than the first in 60.1%. The three-category CD4 classification agreed in 89.1% (kappa 0.82) and the ≤500 cells/µL classification in 93.6%. HIV-RNA ≥5000 copies/mL, present at the first test in 948 people (91.5%), persisted at the second test in 97.6% (95% CI 96.4 to 98.4), and any laboratory criterion persisted in 97.7%. Persistence was lower for haemoglobin (75.3%), white blood cells (56.5%) and platelets (55.2%), but these criteria were met at the first test by only 9.4%, 4.4% and 2.8%.

**Table 4.** Agreement between the first test and the two-test rule for CD4 counts and laboratory criteria (two-test pathway, n=1036)

| Analysis | Comparison | n | Pearson r | ICC(2,1) (95% CI) | Mean difference, cells/μL (95% LoA) | Ratio second or mean/first, geometric mean (95% LoA) | 3-category agreement, % ($\kappa$) | $\leq 500$ vs $> 500$ agreement, % |
| --- | --- | --- | --- | --- | --- | --- | --- | --- |
| Main | First vs mean of two | 1036 | 0.962 | 0.959 (0.952–0.965) | +15.9 (-101 to 133) | 1.06 (0.69–1.62) | 89.1 ($\kappa$ 0.82) | 93.6 |
| Main | First vs second | 1036 | 0.861 | 0.850 (0.825–0.871) | +31.7 (-202 to 265) | 1.07 (0.47–2.44) | 79.8 ($\kappa$ 0.67) | 87.3 |
| Interval 28–90 days | First vs mean of two | 955 | 0.963 | 0.960 (0.953–0.967) | +17.7 (-95 to 130) | 1.06 (0.69–1.62) | 89.3 ($\kappa$ 0.82) | 93.8 |
| Interval 28–90 days | First vs second | 955 | 0.870 | 0.855 (0.829–0.876) | +35.3 (-190 to 260) | 1.08 (0.48–2.43) | 80.2 ($\kappa$ 0.68) | 87.7 |

**B. Laboratory criteria a–d (positive at the first test and again at the second test)**
| Criterion | Positive at first test, n (%) | Positive at both tests, n | Persistence, % (95% CI) | Overall agreement, % | $\kappa$ |
| --- | --- | --- | --- | --- | --- |
| a. WBC $< 3,000/\mu$L | 46 (4.4) | 26 | 56.5 (42.2–69.8) | 90 | 0.29 |
| b. Hb $< 12$ (M) / $< 11$ (F) g/dL | 97 (9.4) | 73 | 75.3 (65.8–82.8) | 94.4 | 0.68 |
| c. Platelets $< 100,000/\mu$L | 29 (2.8) | 16 | 55.2 (37.5–71.6) | 97.6 | 0.55 |
| d. HIV-RNA $\geq 5,000$ copies/mL | 948 (91.5) | 925 | 97.6 (96.4–98.4) | 95.2 | 0.68 |
| Any of a–d | 950 (91.7) | 928 | 97.7 (96.5–98.5) | 97.9 | 0.88 |

For each criterion, overall agreement and $\kappa$ compare the first and second tests. For "Any of a–d", the comparison is between meeting any criterion at the first test and meeting the same criterion at both tests. Missing values (one WBC, one haemoglobin and two platelet counts at the first test) were treated as not meeting the criterion.

**Figure 3.**
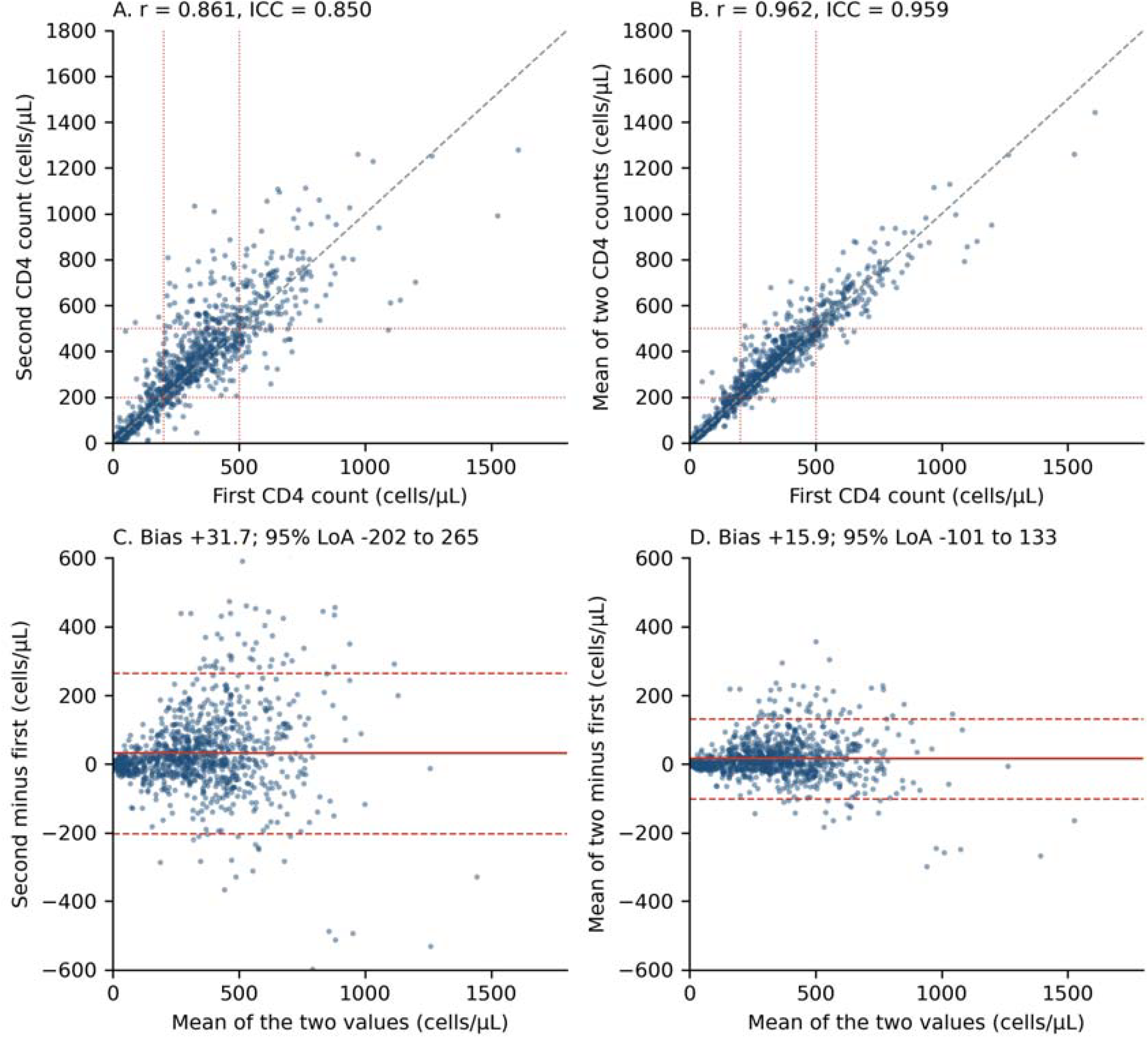
Agreement between the first CD4 count and the two-test rule in the two-test pathway (n = 1036). (A) First versus second CD4 count and (B) first CD4 count versus the mean of the two counts, with the line of identity (dashed) and the 200 and 500 cells/µL thresholds (dotted). (C, D) Bland-Altman plots of the corresponding differences against the mean of the two values; the solid line is the mean difference and the dashed lines are the 95% limits of agreement. ICC, intraclass correlation coefficient; LoA, limits of agreement.

At k ≥1, the two judgements agreed on certification status in 99.0% of people (10 discordant; 0.97%, 95% CI 0.53 to 1.77): six were certifiable on the first test only and four on the two-test rule only (Table 5, Table S4, Figure 4A). Among 112 people whose first CD4 count was 450 to 550 cells/µL, six (5.4%) were classified differently, compared with four of 924 (0.4%) elsewhere, and the odds of discordance halved for every 50 cells/µL that the first count lay from 500 (odds ratio 0.53, 95% CI 0.35 to 0.80; Figure 4D). At k = 0, agreement was 92.5% (78 discordant).

**Table 5.** Concordance of certification status and disability grade between a first-test judgement and the two-test rule across assumed numbers of non-laboratory criteria (two-test pathway, n=1036)

| Assumed number of non-laboratory criteria met (k) | Certified, first test only, n | Certified, two-test rule, n | Certification agreement, % (discordant, n) | Certified by first test only / by two-test rule only, n | Exact grade agreement, % ($\kappa$) | First test gives milder grade, % (one-sided 95% upper bound) | First test gives more severe grade, % |
| --- | --- | --- | --- | --- | --- | --- | --- |
| 0 | 805 | 765 | 92.5 (78) | 59 / 19 | 91.4 (0.78) | 1.9 (2.8) | 6.7 |
| 1 | 1007 | 1005 | 99.0 (10) | 6 / 4 | 94.3 (0.78) | 0.5 (1.0) | 5.2 |
| 2 | 1007 | 1005 | 99.0 (10) | 6 / 4 | 87.5 (0.81) | 2.9 (3.9) | 9.6 |
| 3 | 1007 | 1005 | 99.0 (10) | 6 / 4 | 93.7 (0.87) | 1.3 (2.0) | 5.0 |
| 4 | 1007 | 1005 | 99.0 (10) | 6 / 4 | 90.7 (0.82) | 1.3 (2.0) | 8.0 |
| 5 | 1007 | 1005 | 99.0 (10) | 6 / 4 | 92.7 (0.86) | 1.4 (2.1) | 6.0 |
| 6 | 1007 | 1005 | 99.0 (10) | 6 / 4 | 93.0 (0.87) | 1.4 (2.1) | 5.7 |
| 7 | 1007 | 1005 | 99.0 (10) | 6 / 4 | 93.0 (0.87) | 1.4 (2.1) | 5.7 |
| 8 | 1007 | 1005 | 99.0 (10) | 6 / 4 | 93.0 (0.87) | 1.4 (2.1) | 5.7 |

The non-laboratory criteria (items e–l of the certification table: fatigue, weight loss, fever, diarrhoea, vomiting, opportunistic infections, dietary restrictions, restriction of work) were not collected; the same k was assigned to both judgements and to every person to isolate the contribution of repeat laboratory testing; each row is one such scenario. Certification status differs only between k=0 and k≥1. The one-sided upper bound is the Wilson 95% bound for the proportion in which the first-test judgement gives a milder grade than the two-test rule, compared with a 5% reference threshold. Combining k = 1 to 8, 33 people (3.2%; one-sided 95% upper bound 4.2%) received a milder grade from the first test under at least one k. Missing laboratory values at the first test (four values in two people) were treated as not meeting the criterion; recoding them did not change the number of discordant certification decisions.

**Figure 4.**
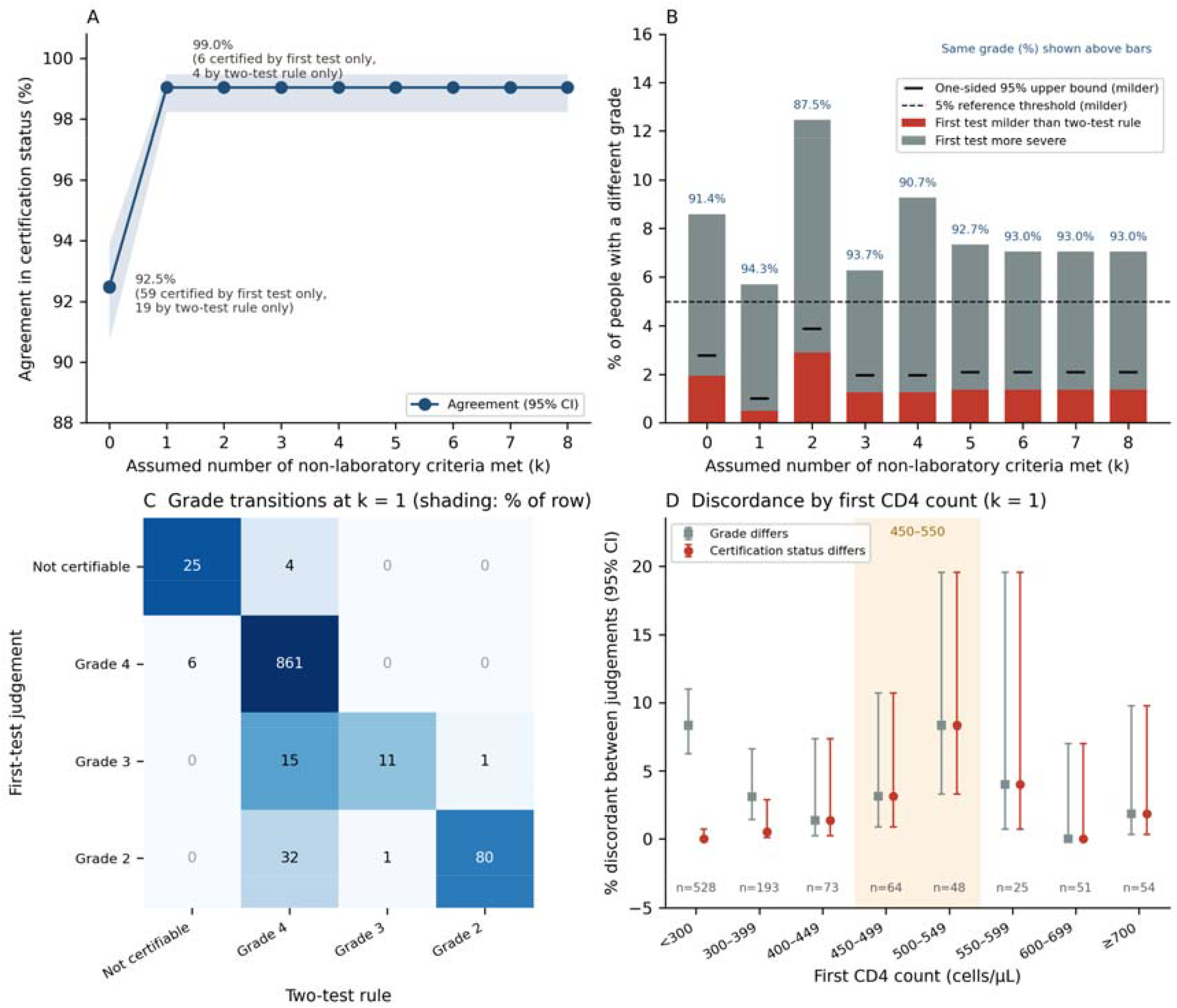
Agreement between the first-test judgement and the two-test rule in the two-test pathway (n = 1036). (A) Agreement in certification status across the assumed number of non-laboratory criteria (k = 0 to 8), with 95% confidence intervals (Wilson); the numbers certifiable on the first test only and on the two-test rule only are given for k = 0 and k = 1. (B) Proportion of people in whom the first test gave a milder (red) or a more severe (grey) grade than the two-test rule, by k; the black ticks are the one-sided 95% upper confidence bounds for the milder proportion, compared with the 5% reference threshold (dashed line); the proportion with the same grade is given above each bar. (C) Cross-tabulation of the grade under the first-test judgement (rows) and the two-test rule (columns) at k = 1; shading shows the percentage of each row, and no one was placed in grade 1. (D) Proportion with a different grade (grey squares) and a different certification status (red circles) at k = 1 by the first CD4 count, with 95% confidence intervals (Wilson); the shaded band marks 450 to 550 cells/µL. CI, confidence interval.

Exact agreement in grade ranged from 87.5% to 94.3% across k = 1 to 8 (91.4% at k = 0). At k = 1, the grade differed in 59 people (5.7%), more severe on the first test in 54 and milder in five (Figure 4C). Most differences (48) arose because a haemoglobin, white blood cell or platelet criterion met at the first test was not met at the second, moving 32 people with a first CD4 count ≤200 cells/µL from grade 2 to 4; only 11 arose from the CD4 count alone. Grade discordance was therefore concentrated among people with low counts (odds ratio 1.16 per 50 cells/µL from 500, 95% CI 1.07 to 1.25). Across all k, the first test gave a milder grade in 0.5% to 2.9% of people (one-sided 95% upper bound at most 3.9%, below the 5% reference threshold) and a more severe grade in 5.0% to 9.6% (Figure 4B). Results were similar in the 955 pairs 28 to 90 days apart (certification agreement 99.2%, eight discordant; Table S1).

### Outcomes after ART

Among 1215 people with post-ART data, HIV-RNA fell below 200 copies/mL after a median of 32 days, in 90.9% by 90 days and 98.0% by one year. Suppression was slower with higher pre-treatment HIV-RNA (adjusted hazard ratio 0.62 per log10, 95% CI 0.57 to 0.68) and with non-integrase inhibitor regimens (0.47, 95% CI 0.30 to 0.75), but did not differ by pathway (Tables S3A to S3C). The median CD4 count at 12 months was 514 cells/µL in the two-test pathway and 269 cells/µL in the single-test pathway; after adjustment for the pre-treatment count, the 12-month count was not associated with time to ART, and the 24-month count was slightly lower with longer waits (−18 cells/µL per log-day, 95% CI −34 to −1; Table S3D). Thirty-nine people (3.0%) died, all after starting ART.

## Discussion

In this cohort of 1302 people newly diagnosed with HIV in Japan, two thirds presented late, and treatment then waited on paperwork. People in the two-test pathway started ART a median of 70 days after their first visit, compared with 11 days for people exempt from the rule, and fewer than 1% started within two weeks. The delay was longest for those with the highest CD4 counts, who are least likely to be certified, and varied threefold between hospitals. The second blood test almost never changed the outcome: HIV-RNA ≥5000 copies/mL persisted in 98% and certification status was identical in 99%. When judgements differed, the first test more often gave a more severe grade, so a single-test rule would rarely disadvantage an applicant.

The 70-day median is largely structural: 28 days is the minimum interval between tests, and the 35-day median between the second test and ART reflects certification processing and scheduling. This contrasts with guidelines that recommend ART within days of diagnosis [1–4] and with the cascade Japan otherwise achieves [13,14], and agrees with earlier Japanese reports [16,17]. The criteria were written when treatment was reserved for advanced disease and still reward a lower CD4 count, so under universal ART the healthiest people wait longest: 3% of the two-test pathway could not be certified and waited a median of 11 months, and a further 16% had a CD4 count >500 cells/µL with a persistent laboratory criterion, most of whom could be certified at grade 4 only with a non-laboratory criterion. Certification is decided by prefectural governments, so variation between facilities may reflect prefectural review as well as hospital practice; our data cannot separate the two. The facility with the shortest intervals started ART on the day of the second draw in 57% of people, and two facilities treated people with CD4 counts >500 cells/µL within two months, so faster treatment is possible under the current rule. Conversely, 42 people with an AIDS-defining illness, 33 at one hospital, went through two tests although the AIDS exemption allowed certification on one; hospitals appear to follow the two-test procedure by default.

The second test adds nothing that the first does not already show. The within-person coefficient of variation of about 26%, which includes the systematic rise between tests, is close to the 25% reported by Hughes et al. in untreated people [22], and discordant decisions clustered near the 500 cells/µL threshold, where single measurements misclassify ART eligibility [26]. The second count was higher than the first, which may reflect transient lowering of circulating CD4 cells by acute illness at diagnosis, so averaging two counts moves people away from certification whereas the first count errs toward it. HIV-RNA ≥5000 copies/mL, present in 92% at diagnosis, was stable and effectively decided eligibility; the haematological criteria fluctuated and accounted for most grade differences, but they were rare and their loss changed the grade, not certification. The two-test interval exists to show that the impairment is fixed, as the disability framework requires, but for HIV this is a formality. Infection is lifelong, and untreated viraemia and CD4 depletion do not resolve except in rare controllers (nine people, 0.7%, had a first HIV-RNA <200 copies/mL), so the diagnosis itself establishes that the impairment will persist. The Ministry’s own guidance recognises this: re-certification is not required while a person remains on ART, even though treatment reverses the very findings the criteria measure [21]. What the certificate certifies, in effect, is the need for lifelong treatment, which one positive test already shows.

The cost of the delay is not primarily immunological: CD4 counts rose while people waited, viral suppression did not differ by pathway, and CD4 recovery at 12 months was unrelated to waiting time. The cost is a longer period of untreated viraemia, during which onward transmission remains possible [11,12], the burden of a diagnosis without treatment, and the risk of disengagement where initiation is slow [8,9].

Our findings support three changes. First, certification could be based on a single test, with the second test replaced by routine monitoring after ART, removing at least four weeks of delay without changing eligibility for 99% of people. Second, ART initiation should be separated from the subsidy procedure, for example by accepting the subsidy application at diagnosis with interim coverage until certification. Third, under universal treatment the CD4 threshold and laboratory criteria no longer identify who needs ART; eligibility based on the diagnosis itself would align the welfare system with treatment guidelines.

This study was retrospective, and the non-laboratory criteria were not recorded. Certification status changed only between zero and one assumed criterion, but we emulated the criteria rather than observing certification decisions, which prefectures may apply differently. The two-test pathway consisted of people for whom clinicians chose to wait, so the agreement estimates apply to that population, and 32 pairs <28 days apart may have been followed by an unrecorded third test. Time to viral suppression depends on testing frequency. Reasons for long waits among people with CD4 counts ≤500 cells/µL were not recorded, and facilities contributed different calendar years. The cohort was 96% male and 86% men who have sex with men, consistent with reported diagnoses in Japan [15,32] but not necessarily representative, and all hospitals were designated AIDS core hospitals, so the findings may not apply to smaller clinics.

## Conclusions

In Japan, the statutory requirement for a second blood test before HIV disability certification adds at least four weeks before ART, delays people with the highest CD4 counts for months, and changes the certification decision in about one person in 100, concentrated near the 500 cells/µL threshold. Certification on a single test, with treatment separated from the subsidy procedure, would let people start ART within days of diagnosis without loss of access to financial support.

## Authors’ contributions

T.T. designed the study, analysed the data and wrote the first draft. T.M., R.M., H.N., E.A., R.S., K.F., J.I., Y.Y., M.S., Y.H. and M.I. collected and verified the data at the participating hospitals and interpreted the results. All authors critically revised the manuscript and have read and approved the final version.

## Acknowledgements

We thank the staff of the participating hospitals who registered and verified the data. This work was supported by Health, Labour and Welfare Sciences Research Grants from the Ministry of Health, Labour and Welfare of Japan (grant numbers 25CA2023A and 26HB1001).

## Data availability statement

Individual-level data cannot be shared publicly because participants did not consent to public release and the data contain sensitive health information. Aggregate data and the analysis scripts, including the emulation of the certification criteria described in the Methods and Supplementary Text S1, are available from the corresponding author on reasonable request.

## Supporting information

Supporting Information file 1 (PDF): Text S1, the Japanese disability certification and medical care subsidy system for HIV; Table S1, certification concordance in pairs with a 28 to 90 day interval; Table S2, initial antiretroviral regimens; Tables S3A to S3D, time to HIV-RNA <200 copies/mL (Kaplan-Meier, Cox model, sensitivity model) and CD4 count after antiretroviral therapy; Table S4, the 10 people whose certification status differed between judgements; Table S5, time to antiretroviral therapy by year of first visit.

## List of abbreviations

AIDS: acquired immunodeficiency syndrome
ART: antiretroviral therapy
CI: confidence interval
HR: hazard ratio
ICC: intraclass correlation coefficient
IQR: interquartile range
WBC: white blood cell

