## Supporting Information for "One test is enough: Japan’s two-test rule for HIV treatment subsidies delays antiretroviral therapy without changing eligibility"

### Text S1. The Japanese disability certification and medical care subsidy system for HIV

#### S1.1 Why certification governs the timing of antiretroviral therapy

Japan has universal health insurance. Outpatient care, including antiretroviral therapy (ART), is reimbursed with a 30% co-payment for most working-age adults, subject to a monthly ceiling under the high-cost medical expense benefit that depends on income (for an average income, roughly 80,000 yen per month plus 1% of costs above a threshold; figures are illustrative and refer to the rules in force during the study period). At an illustrative drug cost of 200,000 yen per month for a modern single-tablet regimen, a 30% co-payment would amount to about 60,000 yen per month before the ceiling is applied, and the ceiling itself would still be a large recurring expense for most households. Almost all people with HIV therefore rely on the medical care subsidy for persons with disabilities (Jiritsu-shien iryo, "self-reliance support medical care", rehabilitation medical care category) under the Act on Comprehensive Support for Persons with Disabilities. The subsidy reduces the co-payment for HIV-related care to 10% and caps it at a monthly amount that depends on household income (currently 0 yen for households receiving public assistance, 2,500 or 5,000 yen for low-income households, 5,000 or 10,000 yen for middle-income households, and 20,000 yen for higher-income households under the "severe and continuous" category that applies to HIV). The subsidy applies from the date the subsidy application is accepted by the municipality and is not paid retroactively for care received before that date; the subsidy application in turn requires the disability certificate, although some municipalities accept it while the certificate application is pending, and the date from which coverage starts and the procedure for simultaneous application vary by prefecture and municipality.

Eligibility for the subsidy requires a physical disability certificate (shintai shogaisha techo) for "immune dysfunction due to human immunodeficiency virus", one of the categories of physical disability defined under the Act on Welfare of Physically Disabled Persons. The category was introduced in 1998. The laboratory requirements of the criteria, including the two-test rule, have not been changed since, although the accompanying guidance has been amended (for example, the 2016 revision of the question on re-certification). A physician designated under Article 15 of the Act completes a standard medical certificate; the person applies through the municipality; the prefecture (or designated city) reviews the certificate and issues the certificate with a grade from 1 (most severe) to 4. The person then applies for the medical care subsidy, which is granted for one year and renewed annually. In a typical sequence, the steps from the first hospital visit to the start of subsidised ART are: first blood test, second blood test at least four weeks later, completion of the medical certificate, municipal application, prefectural review and issuance, subsidy application, and ART. Hospitals differ in whether they start ART at the second blood draw and complete the paperwork in parallel, or wait for the subsidy decision, and municipalities differ in whether the subsidy application can be lodged before the certificate is issued.

#### S1.2 Certification criteria for people aged 13 years or older

The criteria (Ministry of Health, Labour and Welfare notification on the interpretation of the grading table of physical disabilities, 2003, cited here in its current consolidated text) apply to people aged 13 years or older; separate criteria exist for children. They define four grades using the CD4 count, an AIDS-defining illness and the 12 items of Table S1-1.

Table S1-1. Items of the certification table ("Table 1" of the criteria)

| Item | Criterion | Type |
| --- | --- | --- |
| a | White blood cell count <3,000/μL on two consecutive tests at least four weeks apart | Laboratory |
| b | Haemoglobin <12 g/dL (men) or <11 g/dL (women) on two consecutive tests at least four weeks apart | Laboratory |
| c | Platelet count <100,000/μL on two consecutive tests at least four weeks apart | Laboratory |
| d | HIV-RNA ≥5,000 copies/mL on two consecutive tests at least four weeks apart | Laboratory |
| e | Strong fatigue and easy fatigability requiring at least one hour of bed rest per day on seven or more days per month | Non-laboratory |
| f | Weight loss of 10% or more compared with the healthy state | Non-laboratory |
| g | Unexplained fever (≥38°C) on seven or more days per month for two or more months | Non-laboratory |
| h | Loose or watery diarrhoea three or more times a day on seven or more days per month | Non-laboratory |
| i | Vomiting two or more times a day, or nausea lasting 30 minutes or more, on seven or more days per month | Non-laboratory |
| j | History of opportunistic infection (recurrent oral candidiasis, amoebic dysentery, herpes zoster, recurrent herpes simplex, strongyloidiasis, molluscum contagiosum and similar) | Non-laboratory |
| k | Need for restrictions in daily life such as avoidance of raw foods | Non-laboratory |
| l | Need to avoid work beyond light (desk) work | Non-laboratory |

Table S1-2. Grades

| Grade | Criteria (any one of the following) |
| --- | --- |
| 1 | (1) CD4 count ≤200 cells/μL and six or more items of Table S1-1; or (2) irreversible AIDS-related complications making daily life almost impossible without assistance |
| 2 | (1) CD4 count ≤200 cells/μL and three or more items; or (2) a history of AIDS and three or more items; or (3) regardless of CD4 count, six or more items including at least one of items a to d |
| 3 | (1) CD4 count ≤500 cells/μL and three or more items; or (2) regardless of CD4 count, four or more items including at least one of items a to d |
| 4 | (1) CD4 count ≤500 cells/μL and one or more items; or (2) regardless of CD4 count, two or more items including at least one of items a to d |

Notes attached to the criteria specify the following. The CD4 count used for grading is "the lowest value to date of the mean of two consecutive tests at least four weeks apart". Each laboratory item a to d requires the abnormal value to be present on two consecutive tests at least four weeks apart (Table S1-1); the pairs of tests for white blood cells, haemoglobin, platelets and HIV-RNA need not coincide with each other, and the values are taken as "the lowest values to date", which the official questions and answers interpret as the values at the time of the worst state for each item (Section S1.3). A month means 30 consecutive days rather than a calendar month, so that "7 or more days per month" in items e to i means 7 or more days, not necessarily consecutive, within any 30 consecutive days. "Restrictions in daily life" (item k) include, in addition to restriction of raw foods, restriction of untreated water, long-term intensive treatment, strict medication management and avoidance of crowds, and "light work" means desk work. AIDS is diagnosed according to the national surveillance case definition. These notes are set out in the certification procedures guidance (Notice Shoki-hatsu No. 0110001, 10 January 2003, as amended; section 10), which accompanies the criteria (Notice Sho-hatsu No. 0110001, 10 January 2003, as amended).

#### S1.3 Official interpretations relevant to this study

A Ministry of Health, Labour and Welfare notice of questions and answers on the certification criteria (Notice Shoki-hatsu No. 0227001, 27 February 2003, cited in its current consolidated text) contains the following interpretations for immune dysfunction due to HIV.

The purpose of the requirement for two tests at least four weeks apart is "to confirm that the value persists below (or above) the threshold, and thereby to evaluate the impairment of immune function"; an interval of 27 days between monthly visits on a fixed weekday may be regarded as four weeks.

Laboratory values are in principle the most recent values at the time of application regardless of treatment. For a person who has already started ART, however, values obtained before treatment may be used for certification; certifying on hypothetical untreated values is not appropriate.

For items a to d, "the lowest values to date" means that, when several results are available on different dates, the values at the time of the worst state are used for each item; how far apart the individual tests may be is left to the judgement of the certifying physician, with reference to the period in which daily-life restrictions are assessed.

The laboratory items a to d are given greater weight than items e to l because they are objective measurements; with the same number of items, the absence of any laboratory item may lead to a lower grade.

While a person continues ART, re-certification is in principle not required, "in view of the nature of this disability", even though the grade would be expected to change under treatment.

#### S1.4 Implications for the present study

Three features of the system shape the analysis. First, the second blood test is the only element of the criteria that a clinician must wait for: items e to l are symptoms and daily-life restrictions recorded by the physician, and item k, which includes strict medication management, applies to essentially everyone who is about to start lifelong ART. Second, because the CD4 count is averaged and each laboratory item must be met twice, certification status under the two-test rule can differ from a judgement made on the first test through the CD4 count and items a to d; the emulation in the main text therefore holds the number of non-laboratory items constant between the two judgements to isolate the contribution of repeat laboratory testing. Third, the guidance already allows pre-treatment values to be used for people who have started ART and does not require re-certification during treatment, which is consistent with the view that the impairment is established by the diagnosis itself.

### Table S1. Certification concordance restricted to pairs with a 28 to 90 day interval (n=955)

| k | n | Certification agreement, % | Discordant, n | Exact grade agreement, % | First test milder, % (upper bound) |
|---|---|---|---|---|---|
| 0 | 955 | 92.5 | 72 | 91.3 | 1.8 (2.6) |
| 1 | 955 | 99.2 | 8 | 94.2 | 0.3 (0.8) |
| 2 | 955 | 99.2 | 8 | 87.7 | 2.5 (3.5) |
| 3 | 955 | 99.2 | 8 | 93.8 | 1.0 (1.7) |
| 4 | 955 | 99.2 | 8 | 90.8 | 1.0 (1.7) |
| 5 | 955 | 99.2 | 8 | 92.8 | 1.2 (1.9) |
| 6 | 955 | 99.2 | 8 | 93.1 | 1.2 (1.9) |
| 7 | 955 | 99.2 | 8 | 93.1 | 1.2 (1.9) |
| 8 | 955 | 99.2 | 8 | 93.1 | 1.2 (1.9) |

k, assumed number of non-laboratory criteria (items e–l) assigned to every person in the scenario. The one-sided upper bound is the Wilson 95% bound for the proportion in which the first-test judgement gives a milder grade than the two-test rule. The 28 to 90 day window is an analytical restriction, not a regulatory limit.

**Table S2. Initial antiretroviral regimens among people who started antiretroviral therapy (n = 1287): anchor drug**

| **Category** | **n** | **%** |
|---|---|---|
| BIC | 642 | 49.9 |
| DTG | 494 | 38.4 |
| RAL | 75 | 5.8 |
| EVG/c | 50 | 3.9 |
| NNRTI | 17 | 1.3 |
| PI | 9 | 0.7 |
| Other/unclassified | 0 | 0.0 |

Nucleoside backbone

| **Category** | **n** | **%** |
|---|---|---|
| TAF/FTC | 954 | 74.1 |
| ABC/3TC | 170 | 13.2 |
| TDF/FTC | 125 | 9.7 |
| 3TC-only | 25 | 1.9 |
| other | 11 | 0.9 |
| ISL | 2 | 0.2 |
| non_regimen | 0 | 0.0 |

Regimen characteristics

| **Item** | **n** | **%** |
|---|---|---|
| Single-tablet regimen | 822 | 63.9 |
| Boosted regimen (ritonavir or cobicistat) | 59 | 4.6 |
| Two-drug regimen (two antiretroviral agents) | 33 | 2.6 |
| Four or more antiretroviral agents (boosters excluded) | 10 | 0.8 |
| Four or more components (boosters included) | 65 | 5.1 |
| Unclassified | 0 | 0.0 |

Most frequent regimens

| Regimen | n | % |
|---|---|---|
| BIC/TAF/FTC | 639 | 49.7 |
| DTG+TAF/FTC | 216 | 16.8 |
| ABC/3TC/DTG | 130 | 10.1 |
| DTG+TDF/FTC | 81 | 6.3 |
| RAL+TAF/FTC | 42 | 3.3 |
| EVG/COBI/TAF/FTC | 28 | 2.2 |
| DTG+ABC/3TC | 22 | 1.7 |
| DTG/3TC | 20 | 1.6 |
| RAL+TDF/FTC | 17 | 1.3 |
| EVG/COBI/TDF/FTC | 13 | 1.0 |
| DOR+TAF/FTC | 7 | 0.5 |
| ABC/3TC+DTG | 7 | 0.5 |
| RAL+ABC/3TC | 7 | 0.5 |
| RAL+DOR | 6 | 0.5 |
| RPV+TAF/FTC | 4 | 0.3 |

Denominator for all percentages: 1287 people who started antiretroviral therapy. Regimen characteristics are overlapping attributes, not mutually exclusive categories. Antiretroviral agents were counted from the recorded regimen components; the pharmacokinetic boosters ritonavir and cobicistat are excluded from the agent count and reported separately, and the number of components including boosters is shown for comparison. Regimens are classified by drug components after harmonising recorded spellings; co-formulated and separately administered combinations of the same components are shown as recorded in the source table. BIC, bictegravir; DTG, dolutegravir; RAL, raltegravir; EVG/c, elvitegravir/cobicistat; NNRTI, non-nucleoside reverse transcriptase inhibitor; PI, protease inhibitor.

**Table S3A. Time from antiretroviral therapy initiation to HIV-RNA <200 copies/mL (Kaplan-Meier), excluding one hospital without post-treatment data**

| Stratum | n | Events | Median days | % by 90 days | % by 180 days | % by 365 days |
|---|---|---|---|---|---|---|
| All | 1215 | 1178 | 32.0 | 90.9 | 96.9 | 98.0 |
| Anchor: BIC | 588 | 570 | 31.0 | 91.0 | 96.5 | 98.1 |
| Anchor: DTG | 483 | 472 | 32.0 | 92.0 | 98.2 | 98.7 |
| Anchor: RAL | 73 | 68 | 44.0 | 86.1 | 93.0 | 93.0 |
| Anchor: EVG/c | 50 | 48 | 28.0 | 90.7 | 97.7 | 97.7 |
| Anchor: NNRTI | 13 | 12 | 49.0 | 66.7 | 83.3 | 91.7 |
| Anchor: PI | 8 | 8 | 70.0 | 100.0 | 100.0 | 100.0 |
| Backbone: TAF/FTC | 892 | 864 | 34.0 | 90.8 | 96.8 | 98.2 |
| Backbone: TDF/FTC | 122 | 116 | 34.0 | 85.7 | 95.0 | 96.0 |
| Backbone: ABC/3TC | 169 | 167 | 28.0 | 95.2 | 98.8 | 98.8 |
| Backbone: 3TC-only | 19 | 19 | 34.0 | 94.7 | 100.0 | 100.0 |
| Baseline HIV-RNA $\leq 10^4$ | 171 | 166 | 28.0 | 97.3 | 98.6 | 98.6 |
| Baseline HIV-RNA $>10^4$–$10^5$ | 578 | 568 | 28.0 | 95.1 | 98.6 | 98.8 |
| Baseline HIV-RNA $>10^5$–$10^6$ | 395 | 380 | 43.0 | 87.8 | 95.7 | 97.0 |
| Baseline HIV-RNA $>10^6$ | 71 | 64 | 78.0 | 59.2 | 85.7 | 96.3 |
| Baseline CD4 ≤200 | 479 | 454 | 45.0 | 83.7 | 94.1 | 97.0 |
| Baseline CD4 201–350 | 262 | 259 | 28.0 | 95.5 | 99.2 | 99.2 |
| Baseline CD4 351–500 | 244 | 239 | 28.0 | 93.8 | 97.9 | 97.9 |
| Baseline CD4 >500 | 230 | 226 | 28.0 | 97.8 | 99.1 | 99.1 |
| Two-test pathway | 973 | 951 | 29.0 | 93.1 | 98.0 | 98.4 |
| Single-test pathway | 242 | 227 | 48.0 | 81.9 | 92.5 | 96.4 |
| Two-test, first→ART ≤30 d | 111 | 109 | 35.0 | 91.8 | 98.9 | 100.0 |
| Two-test, first→ART 31–90 d | 530 | 518 | 31.0 | 91.9 | 97.4 | 98.0 |
| Two-test, first→ART 91–180 d | 226 | 224 | 28.0 | 96.4 | 99.6 | 99.6 |
| Two-test, first→ART >180 d | 106 | 100 | 32.0 | 93.4 | 96.7 | 96.7 |

| Stratum | n | Events | Median days | % by 90 days | % by 180 days | % by 365 days |
|---|---|---|---|---|---|---|
| ART started 2015 | 82 | 77 | 29.0 | 88.4 | 94.8 | 94.8 |
| ART started 2016 | 106 | 102 | 28.0 | 91.3 | 97.1 | 97.1 |
| ART started 2017 | 113 | 111 | 31.0 | 91.2 | 99.0 | 99.0 |
| ART started 2018 | 145 | 141 | 30.0 | 95.7 | 98.6 | 98.6 |
| ART started 2019 | 202 | 197 | 34.0 | 92.7 | 97.9 | 97.9 |
| ART started 2020 | 184 | 179 | 32.0 | 88.8 | 97.4 | 98.1 |
| ART started 2021 | 134 | 132 | 29.0 | 93.9 | 97.7 | 98.5 |
| ART started 2022 | 91 | 87 | 41.0 | 86.2 | 97.7 | 98.9 |
| ART started 2023 | 93 | 91 | 35.0 | 89.1 | 93.5 | 98.9 |
| ART started 2024 | 60 | 56 | 35.0 | 84.5 | 89.6 | 96.5 |
| ART started 2025 | 5 | 5 | 35.0 | 100.0 | 100.0 | 100.0 |

Analysis set: 1215 of the 1287 people who started antiretroviral therapy (ART), after excluding 71 at one hospital that did not record post-treatment outcomes and one person who started ART after the fallback censoring date with no follow-up recorded. The event is the first recorded HIV-RNA <200 copies/mL; people without a recorded event were censored at the recorded end of follow-up or, when none was recorded (n = 12), at 30 June 2025. Pre-treatment (baseline) HIV-RNA and CD4 count are the values of the second test in the two-test pathway and of the first test in the single-test pathway. Time to suppression depends on the frequency of HIV-RNA testing, which was not standardised across hospitals.

### Table S3B. Cox model for time to HIV-RNA <200 copies/mL, stratified by facility (reference anchor drug: bictegravir)

| Variable | HR | Lower 95% CI | Upper 95% CI | p |
|---|---|---|---|---|
| Baseline HIV-RNA, per log10 | 0.624 | 0.573 | 0.679 | <0.001 |
| Baseline CD4, per 100 cells/µL | 1.023 | 0.992 | 1.055 | 0.155 |
| Age, per 10 years | 1.005 | 0.951 | 1.062 | 0.864 |
| Male sex | 0.822 | 0.595 | 1.136 | 0.235 |
| AIDS-defining illness | 0.838 | 0.688 | 1.020 | 0.078 |
| Two-test pathway | 1.112 | 0.902 | 1.369 | 0.320 |
| Year of ART start, per year | 0.997 | 0.965 | 1.030 | 0.860 |
| Anchor DTG (vs BIC) | 1.034 | 0.878 | 1.218 | 0.688 |
| Anchor EVG/c (vs BIC) | 1.103 | 0.783 | 1.555 | 0.574 |
| Anchor NNRTI or PI (vs BIC) | 0.472 | 0.295 | 0.754 | 0.002 |

| **Variable** | **HR** | **Lower 95% CI** | **Upper 95% CI** | **p** |
|---|---|---|---|---|
| Anchor RAL (vs BIC) | 0.814 | 0.615 | 1.078 | 0.150 |

n = 1215, events = 1178. Hazard ratios (HR) >1 indicate faster suppression. The model is stratified by facility and adjusted for the variables shown; baseline values are defined as in Table S3A. Proportional hazards were examined with Schoenfeld residuals (rank transformation): the assumption was not met for baseline HIV-RNA ($p < 0.001$), and there was some evidence of non-proportionality for AIDS-defining illness ($p = 0.03$) and the NNRTI/PI group ($p = 0.05$); the HRs for these variables are therefore averages over follow-up, and a model stratified by HIV-RNA category is shown in Table S3C. The time from the first visit to ART was not included as a covariate in this model; its association with CD4 recovery is examined in Table S3D.

**Table S3C. Sensitivity analysis: Cox model for time to HIV-RNA <200 copies/mL stratified by facility and by baseline HIV-RNA category (≤$10^4$, >$10^4$ to $10^5$, >$10^5$ to $10^6$, >$10^6$ copies/mL)**

| Variable | HR | Lower 95% CI | Upper 95% CI | p |
|---|---|---|---|---|
| Baseline CD4, per 100 cells/μL | 1.032 | 0.999 | 1.065 | 0.056 |
| Age, per 10 years | 1.000 | 0.945 | 1.058 | 0.992 |
| Male sex | 0.717 | 0.516 | 0.996 | 0.047 |
| AIDS-defining illness | 0.865 | 0.704 | 1.063 | 0.167 |
| Two-test pathway | 1.165 | 0.932 | 1.456 | 0.180 |
| Year of ART start, per year | 0.990 | 0.956 | 1.025 | 0.573 |
| Anchor DTG (vs BIC) | 0.980 | 0.827 | 1.162 | 0.816 |
| Anchor EVG/c (vs BIC) | 0.977 | 0.688 | 1.389 | 0.899 |
| Anchor NNRTI or PI (vs BIC) | 0.434 | 0.262 | 0.719 | 0.001 |
| Anchor RAL (vs BIC) | 0.766 | 0.573 | 1.024 | 0.072 |

n = 1215, events = 1178. Same covariates as Table S3B except baseline HIV-RNA, which is used as a stratification variable (categories are right-inclusive: ≤$10^4$, >$10^4$ to $10^5$, >$10^5$ to $10^6$, >$10^6$ copies/mL; the same categories are used in Table S3A). In this model no covariate showed evidence of non-proportional hazards (Schoenfeld residual tests, all $p \geq 0.05$; AIDS-defining illness $p = 0.05$).

**Table S3D. CD4 count after antiretroviral therapy by testing pathway (median cells/μL and % ≥500 at each time point)**

| Stratum | n | Pre-ART median | 12 mo n | 12 mo median | 12 mo % ≥500 | 24 mo n | 24 mo median | 24 mo % ≥500 | 36 mo n | 36 mo median | 36 mo % ≥500 | 48 mo n | 48 mo median | 48 mo % ≥500 | 60 mo n | 60 mo median | 60 mo % ≥500 |
|---|---|---|---|---|---|---|---|---|---|---|---|---|---|---|---|---|---|
| Two-test pathway | 974 | 330 | 840 | 514 | 53 | 789 | 548 | 58 | 694 | 572 | 62 | 624 | 564 | 63 | 525 | 581 | 65 |
| Single-test pathway (AIDS) | 196 | 34 | 157 | 242 | 7 | 148 | 313 | 16 | 134 | 328 | 24 | 113 | 344 | 26 | 102 | 366 | 32 |
| Single-test pathway (other) | 29 | 232 | 22 | 508 | 50 | 24 | 482 | 46 | 22 | 502 | 50 | 18 | 480 | 44 | 13 | 485 | 46 |
| Single-test pathway (acute infection) | 14 | 282 | 13 | 602 | 77 | 13 | 716 | 77 | 13 | 778 | 77 | 10 | 836 | 80 | 8 | 814 | 75 |
| Single-test pathway (expected HIV-RNA ≥5000) | 3 | 41 | 3 | 266 | 0 | 2 | 392 | 0 | 2 | 410 | 0 | 2 | 454 | 50 | 1 | 519 | 100 |

Analysis set: 1216 people who started ART, excluding 71 at one hospital that did not record post-treatment CD4 counts. At each time point, n is the number of people with a recorded value, and the median and the percentage ≥500 cells/μL are calculated among them only; values were not imputed, and people who had died, transferred, been lost or not yet reached the time point are not counted as <500. Values were extracted at the scheduled time point (12, 24, 36, 48 and 60 months) as recorded by each hospital. Baseline CD4 is defined as in Table S3A. In linear regression adjusted for baseline CD4 count, baseline HIV-RNA, age, sex, AIDS-defining illness, pathway, anchor drug and facility, the natural logarithm of days from the first visit to ART (values <1 set to 1) was not associated with the CD4 count at 12 months (coefficient -6.3 cells/μL per log-day, 95% CI -19.7 to 7.1, p = 0.36; n = 1035) and was weakly associated at 24 months (-17.5, 95% CI -34.1 to -1.0, p = 0.038; n = 976); these coefficients are per unit of log-days, not per day.

### Table S4. The 10 people whose certification status differed between the first-test judgement and the two-test rule (k=1)

| Case | Facility | First CD4 | Second CD4 | Mean CD4 | First HIV-RNA, log10 | Second HIV-RNA, log10 | Laboratory criteria first / both | Grade, first test (k=1) | Grade, two-test rule (k=1) | First test to ART, days |
|---|---|---|---|---|---|---|---|---|---|---|
| 1 | F | 338 | 792 | 565 | recorded as 0 | 2.04 | 0/0 | 4 | 0 | 146 |
| 2 | F | 400 | 804 | 602 | 3.30 | 3.46 | 0/0 | 4 | 0 | 102 |
| 3 | B | 505 | 472 | 488 | 2.81 | 3.43 | 0/0 | 0 | 4 | 96 |
| 4 | D | 561 | 481 | 521 | 3.74 | 3.45 | 2/0 | 4 | 0 | 77 |
| 5 | G | 475 | 535 | 505 | 3.56 | 3.67 | 0/0 | 4 | 0 | 175 |
| 6 | C | 851 | 986 | 918 | 4.15 | 3.68 | 1/0 | 4 | 0 | 379 |
| 7 | A | 515 | 321 | 418 | 3.18 | 3.30 | 0/0 | 0 | 4 | 172 |
| 8 | A | 503 | 470 | 486 | 2.18 | 2.49 | 0/0 | 0 | 4 | 297 |
| 9 | A | 536 | 464 | 500 | 3.15 | 2.88 | 0/0 | 0 | 4 | 413 |
| 10 | H | 486 | 557 | 522 | 3.57 | 3.20 | 0/0 | 4 | 0 | 666 |

Grade 0 denotes not certifiable (not a statutory grade). "Laboratory criteria first / both" gives the number of items a–d met at the first test and the number met at both tests. One first HIV-RNA value was recorded as 0 copies/mL and is shown as recorded. Mean CD4 counts are rounded for display; grades were assigned from unrounded values.

### Table S5. Time to antiretroviral therapy by year of first test

| Year of first test | n (started ART) | Two-test pathway, % | First test to ART, all, days, median (IQR) | First test to ART, two-test pathway | First test to ART, single-test pathway | Second test to ART, two-test pathway | ART within 14 days of first test, % | Facilities contributing, n |
|---|---|---|---|---|---|---|---|---|
| 2015 | 126 | 84.1 | 77 (39–133) | 106 (52–172) | 24 (13–29) | 56 (21–104) | 4.8 | 7 |
| 2016 | 93 | 84.9 | 70 (39–112) | 75 (50–119) | 18 (4–26) | 42 (14–86) | 6.5 | 6 |
| 2017 | 115 | 81.7 | 70 (28–108) | 78 (52–127) | 15 (9–20) | 42 (14–84) | 8.7 | 6 |
| 2018 | 164 | 75.6 | 68 (28–98) | 77 (58–106) | 8 (4–16) | 45 (22–73) | 15.9 | 7 |
| 2019 | 207 | 82.1 | 56 (29–84) | 63 (39–94) | 7 (3–16) | 33 (7–56) | 12.6 | 7 |
| 2020 | 170 | 77.1 | 50 (28–90) | 63 (41–96) | 10 (4–21) | 33 (8–63) | 14.7 | 8 |
| 2021 | 131 | 82.4 | 54 (29–82) | 64 (36–91) | 8 (4–16) | 35 (7–60) | 13.0 | 8 |
| 2022 | 100 | 76.0 | 42 (28–75) | 56 (37–98) | 6 (2–14) | 21 (7–61) | 20.0 | 8 |
| 2023 | 110 | 76.4 | 49 (28–77) | 62 (39–86) | 12 (4–22) | 28 (9–49) | 14.5 | 7 |
| 2024 | 71 | 76.1 | 49 (30–77) | 60 (42–88) | 14 (12–24) | 28 (8–54) | 12.7 | 7 |

Restricted to people who started ART (n=1287). Intervals are in days, median (IQR). The percentage within 14 days is the observed proportion among ART starters in that year and is not a Kaplan–Meier estimate. The number of contributing facilities varies by year.
